# CLUSTER ANALYSIS IDENTIFIES LONG COVID SUBTYPES IN BELGIAN PATIENTS

**DOI:** 10.1101/2023.11.22.23298807

**Authors:** Pamela Mfouth Kemajou, Tatiana Besse-Hammer, Claire Lebouc, Yves Coppieters

## Abstract

SARS CoV-2 infection presents complications known as long Covid, a multisystemic organ disease which allow multidimensional analysis.

**Objectives:** This study aims to identify Long Covid clusters and to relate them to the clinical classification devised at the Clinical Research Unit of Brugmann University Hospital, Brussels.

**Method:** A two-stage multidimensional exploratory analysis was performed on a cohort of 205 long Covid patients, involving a Factorial Analysis of Mixed Data (FAMD), and then Hierarchical Clustering Post Component Analysis (HCPC).

**Results:** The study’s sample comprised 76% women, with an average age of 44.5 years. Three clinical forms were identified: long, persistent, and post-viral syndrome. Multidimensional analysis identified three clusters: cluster 1 (myalgia-like pain) associated with the persistent clinical form; cluster 2 (neurocognitive disorders) linked to the long clinical form; cluster 3 (neurocognitive disorders, anxio-depressive syndrome, joint pain and myalgia, peripheral nervous system disorders with dysautonomia, including Postural Orthostatic Tachycardia Syndrome, along with digestive system disorders). However, biological data did not provide sufficient differentiation between the clusters.

**Conclusion:** Long Covid phenotypes, as well as clinical forms, appear to be associated with distinct pathophysiological mechanisms or genetic predisposition, warranting further investigation.

## Introduction

Covid-19, caused by the severe acute respiratory syndrome coronavirus, SARS-CoV-2, is an emerging infectious disease. Like SARS-CoV-1, its predecessor, which emerged in 2003 [1], SARS-CoV-2 infection can lead to persistent complications affecting various organ systems collectively referred to as Post Covid Syndrome or PASC (Post Acute Sequelae of SARS-CoV-2) or long Haulers Covid or long Covid [2] [3]. These complications impact approximately 10 to 20% of Covid-19 patients, as defined by the World Health Organization (WHO), which characterizes long Covid as " the continuation or development of new symptoms 3 months after the initial SARS-CoV-2 infection, with these symptoms lasting for at least 2 months with no other explanation"[4].

Long Covid encompasses a spectrum of multisystemic alterations, with various pathophysiological mechanisms that overlap, impairing the development of a comprehensive treatment approach [5] [6] [7]. While numerous symptoms associated with long Covid have been described, none are specific to the disease, but all can be debilitating, especially when they persist for over a year [8].

The distinct timing of symptom onset has therefore led to the establishment of a clinical classification, with three major syndromes recognized at the Clinical Research Unit (CRU) of Brugmann University Hospital, Belgium. This clinical classification include:

- Hypoxemic long Covid patients who require oxygen therapy and often intensive care due to respiratory failure necessitating intubation. Their oxygen saturation levels were, at least on one occasion, less than 89% on ambient air and at rest. Nonetheless, some healthcare professionnals equate this form with PICS (Post Intensive Care Syndrome) and the complications that arise from being hospitalized in an ICU (Intensive Care Unit) [9].
- Persistent forms encompass patients who, following an initial episode of influenza-like illness, experienced multi-organ symptoms that worsened (with the sensation of multiple relapses) from 4 to 8 weeks after infection.
-The "long" forms encompass patients who, within an interval of 1 to 4 weeks, predominantly developed neurocognitive symptoms, followed by multi-organ symptoms, organ by organ. It appears that each "recurrence" led to new symptomatology and exacerbated existing symptoms.

In addition to these three syndromes, two other syndromes are recognized in the literature, namely Mast Cell Activation Syndrome and post-viral syndrome [10] [11]:

-The post-viral or post-Covid syndrome category represents individuals who do not fit into any of the three primary clinical forms aforementioned.
-The Mast Cell Activation Syndrome (MCAS), which primarily presents with digestive symptoms and is diagnosed through tryptase assays [10]. The clinical presentation of MCAS can also involve the cardiopulmonary, gastrointestinal, dermatological, and neurological systems. It is known that long Covid can trigger Mast Cell Activation Syndrome [12].

Beyond this clinical classification, various statistical methods including unsupervised machine learning an clustering have been used to categorize long Covid patients into clusters [13] [14] [15]. However, some of these studies did not consider biological criteria, and others overlooked common clinical signs, such as dysautonomia manifested by exercise intolerance, dyspnoea, and Postural Orthostatic Tachycardia Syndrome (POTS). In this study, we employ comprehensive clinical and biological criteria to identify distinct clusters of long Covid more precisely and establish connections with the aforementioned clinical forms.

## Methodology

### Study design and participants

A retrospective study of medical reports was initiated from January to May 2023 at Brugmann University Hospital. After the medical examination and anonymisation of patient records, only the principal investigator could identify the study participants individually. Collected anonymised data were then coded for analysis purpose and to preserve confidentiality and security of the participants.

Medical records were provided from a cohort study initiated in May 2020 at Brugmann University Hospital to longitudinally monitor patients with a probable or confirmed Covid-19 diagnosis whose symptoms persisted and worsened for more than a month or who developed new symptomatology, even though tests did not reveal the presence of SARS CoV-2 infection or another condition.

### Inclusion and Exclusion Criteria

The study included individuals aged 18 or older who had a probable or confirmed diagnosis of Covid-19, and who had been clinically, radiologically, and neuropsychologically diagnosed with long Covid. Participation in the study was contingent upon the individuals’ voluntary and informed consent. Individuals who had been previously diagnosed with Alzheimer’s disease, demyelinating pathology, dementia, or Mild Cognitive Impairment (MCI), or those with 70% missing information in their records, were excluded from the study.

A total of 281 patients were recruited through May 2023. However, 21 individuals were excluded from the study as they had conditions other than Post Acute Sequelae of SARS-CoV-2 (PASC). Among the remaining 260 patients, 206 were included in our study, while 54 were excluded due to insufficient data. Additionally, an outlier was identified during factorial analysis and subsequently excluded (see figure 1).

**Figure 1:**
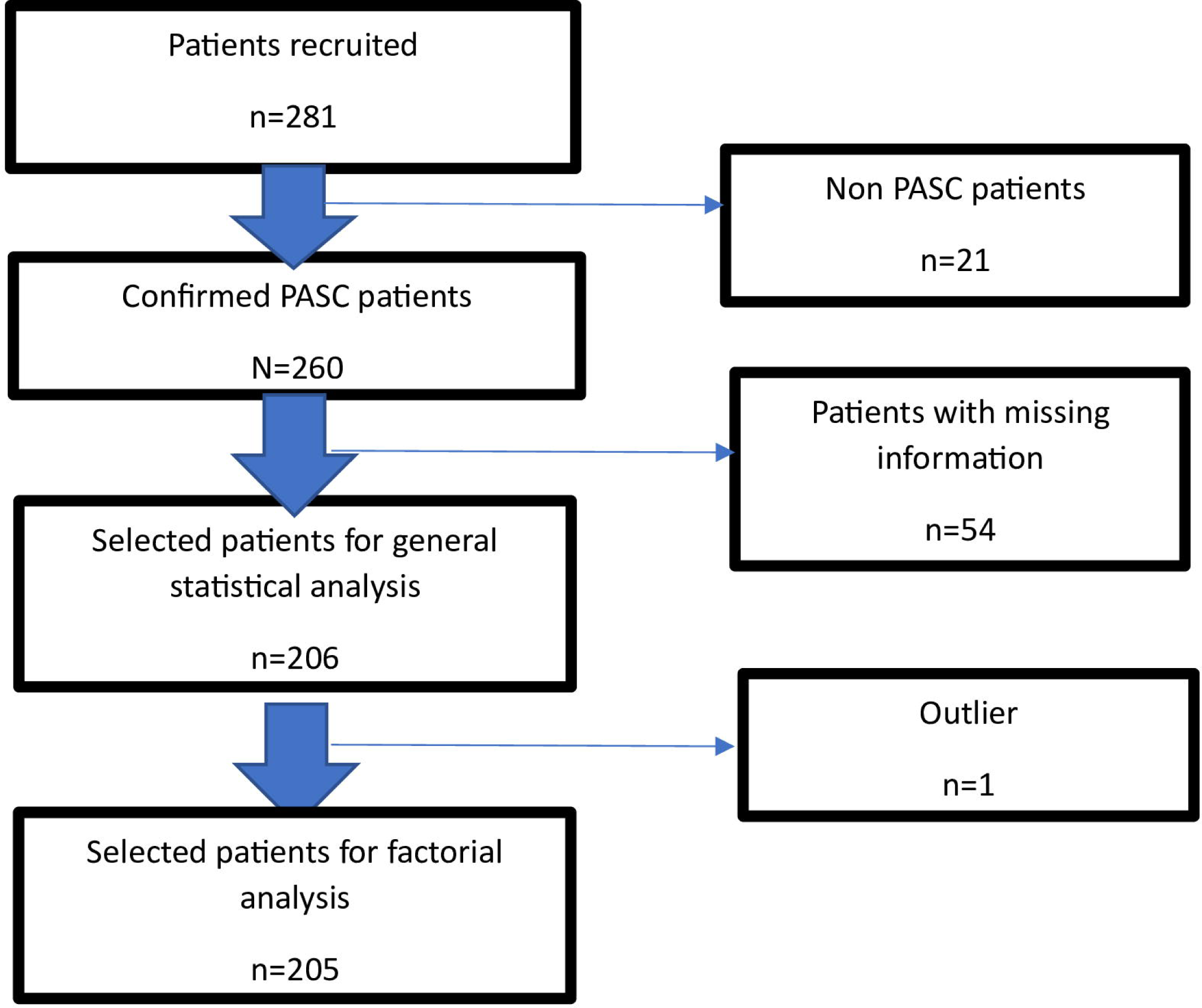
Patient file selection flow chart.

### Ethics

This study was subject to prior approval by the Ethics Committee of the Brugmann University Hospital (ref CE 2022/56) to comply with the principles of the Declaration of Helsinki. Informed consent was required from participants prior to inclusion in the study and before collecting data. The data collected were coded to preserve data confidentiality and patient safety.

### Clinical assessments

Upon obtaining their voluntary consent, participants underwent a comprehensive medical history collection, and a meticulous clinical examination was conducted to identify clinical signs of long Covid, particularly dysautonomia and psychiatric disorders (anxiety and/or depression).

For patients without psychiatric disorders, the following assessments were conducted: Nijmegen tests to diagnose respiratory hyperventilation, the 5-word test to diagnose cortico-subcortical pathology, the MacNair test to screen for neurocognitive disorders, a specific MacNair test to evaluate memory and executive functions.

Symptoms were documented in chronological order, along with their duration, during the inclusion phase, and at 6 and 12 months thereafter. Data pertaining to various biological tests were also collected, which were performed in the post-Covid period and brought in by the patients. If necessary, additional samples were collected to further assess endothelial dysfunction later.

Following the clinical assessment during the inclusion phase, neurocognitive testing was conducted to corroborate the patients’ subjective complaints. If neurocognitive testing yielded positive results, patients underwent brain scintigraphy followed by neurocognitive rehabilitation. In cases where neurocognitive testing was negative, patients were recontacted one year later to monitor changes in their symptoms.

For patients with psychiatric disorders, specifically anxiety and/or depression, a general questionnaire was administered to determine whether the Covid period had induced any stress. The following tests were then carried out: HADS (Hospital Anxiety and Depression Scale) for assessing anxiety and depression, PCL-S (Post-traumatic Checklist Scale) to diagnose PTSD (Post-traumatic Stress Disorder), CFQ (Cognitive Failure Questionnaire) for diagnosing cognitive disorders, SSS-8 (Eight-item Somatic Symptom Scale) for diagnosing complaints related to fatigue.

If any of these four scales indicated abnormalities, patients were referred for psychiatric consultation and treatment. If neurocognitive remediation was deemed necessary, neurocognitive testing was conducted prior to the treatment. Subjective complaints were longitudinally monitored at 6, 12, and 24 months. If none of these scales indicated abnormalities, patients were reevaluated after 22-26 weeks, and the screening scales were repeated.

Psychiatric referrals to various clinics were made based on the following scales: STAI Y-B (State Trait Anxiety Inventory form Y-B) for anxiety traits, Beck for depression, Impact of Event Scale to measure stress perception related to a traumatic event, CTQ (Childhood Trauma Questionnaire), Resilience scale, SF-36 (Short-Form 36) quality of life scale, MOCA (Montreal Cognitive Assessment), PCL-S (Post Traumatic Checklist Scale), FSS (Fatigue Severity Scale), VAS (Visual Analog Scale), DN4 (neurological pain assessment scale), Body chart, Modified Borg scale to assess the perceived intensity of effort during exercise (e.g., a 6-minute walking test).

### General Statistical Analysis

Descriptive analyses were conducted to assess the distribution of various variables. Normally distributed continuous variables were summarized using the mean and standard deviation, while non-normally distributed variables were summarized using the median and interquartile range (IQR). Categorical variables were presented as frequencies. A bivariate analysis was performed to identify variables that were correlated with each other. The Cramer’s V test was used to examine the relationship between two qualitative variables, while Pearson’s linear correlation coefficient was used to investigate the relationship between two quantitative variables. If a variable exhibited a high correlation with another, one of them was excluded from the multivariate analysis.

### Multidimensional Analysis

An exploratory analytical approach utilized two complementary statistical techniques known as multidimensional data analysis methods. These techniques facilitated the creation of synthetic representations of objects (such as individuals, variables, and modalities of qualitative variables) in the form of point clouds within a Euclidean space, which could be reduced to a two-dimensional plane. The Euclidean aspect meant that distances between points and angles for quantitative variables were interpreted in terms of correlation for quantitative variables and similarity for individuals and modalities.

#### Factor Analysis of Mixed Data (FAMD)

FAMD is a principal component analysis method that combines the transformation of categorical data into coordinates in a multidimensional space, with quantitative data transformation. It aims to assess linear relationships between variables by detecting the main dimensions of variability and summarizing them into synthetic variables, thereby reducing the number of variables needed to represent the data.

Before conducting FAMD, several steps were taken:

- Bivariate analysis was performed to exclude highly correlated variables. For instance, LDL cholesterol and total cholesterol levels were highly correlated, leading to the exclusion of the total cholesterol variable from the analysis.
- Active variables (clinical and biological) were distinguished from additional variables that did not contribute to calculating distances between individuals. These additional variables were age categories, gender, wave of SARS-CoV-2 infection, and long Covid clinical form. They were considered explanatory variables but did not participate in constructing the axes.
- A balanced distribution of modalities was ensured to avoid bias in the factorial analysis. Variables with a frequency of less than 10% were excluded from the active variables since they could not be further subdivided into multiple modalities.
- Missing data were imputed by estimating the value of variable x from that of y if they were correlated, or by estimating the value of x from an individual with similarities to it.

During the factorial analysis, an outlier was detected, which had a disproportionate influence on constructing the axes and was subsequently removed from the analysis [16].

#### Cluster analysis or HCPC (Hierarchical Clustering Post Component analysis)

Secondly, cluster analysis or HCPC was used on the coordinates of the mixed analysis using aggregation by inertia to obtain clusters or a partition of the data. This method starts with each participant as its own group, combining the most "similar" participants based on proximity of Euclidean distance, continuing until the last groups merge into 1 group containing all participants: this is Ward’s method. The clusters were then consolidated to make them homogeneous. The clusters were compared using the χ^2^ test for categorical variables and the Kruskall Wallis test for continuous variables.

## Results

### Study Participants

Participants were recruited, on average, 328 ± 176 days (approximately 47 weeks) after the onset of long Covid. The average duration from infection to the onset of long Covid was 65 ± 42.9 days (approximately 9 weeks). The mean age at the diagnosis of long Covid was 44.5 years, and patients presented with an average of 6 symptoms at the time of Covid-19 diagnosis. More than two-thirds of the study population were women, primarily affected during the second wave. Close to half of the participants had the long form of long Covid, and about 10% of all cases required hospitalization (see table 1).

**Table 1:**
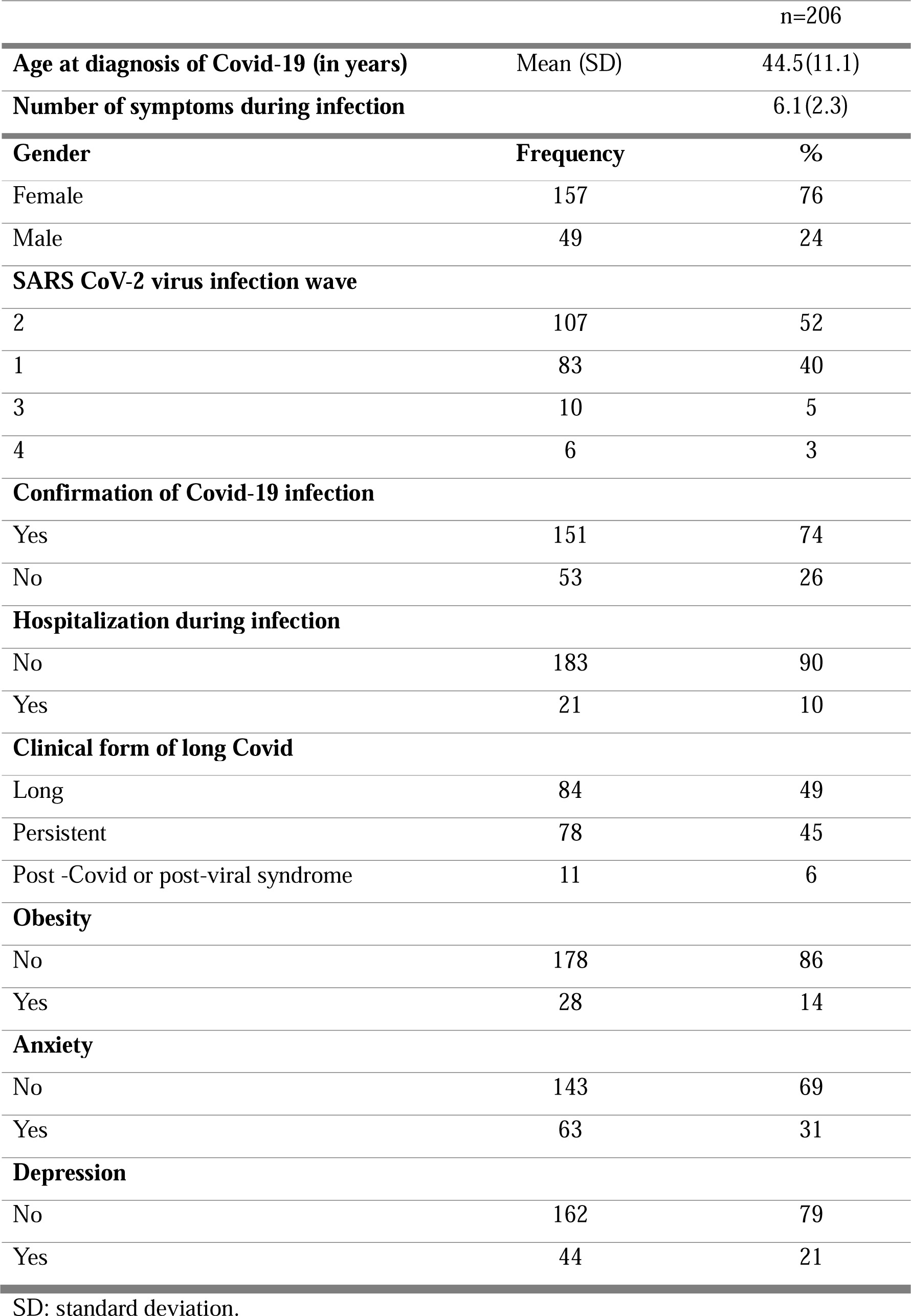
Socio-demographic characteristics of the study population.

### Description of clinical and biological variables

During the clinical examination, 28 primary symptoms and/or syndromes were identified. The most prevalent symptoms of long Covid at inclusion were concentration disorders, memory disorders, fatigue, phasic disorders, and sleep disorders.

Three variables were excluded from the factorial analysis due to missing data: d-dimers, creatine kinase, and lactic dehydrogenase. All biological values fell within the normal ranges except for elevated C-reactive protein (CRP), LDL cholesterol, and total cholesterol. The median CRP was 22 [6-45] mg/L, the mean total cholesterol was 202.5 ± 40.6 mg/dL and the mean LDL cholesterol was 117.9 ± 35.9 mg/ dL.

### Partition of individuals

The individuals, variables, and modalities were represented in a multidimensional space using Factor Analysis of Mixed Data (FAMD), conducted with the Factominer and Factoextra packages of R software version 4.2.

The Hierarchical Clustering Post Component Analysis (HCPC) was then performed after determining the optimal number of principal components to be analyzed. Ideally, 80% of all components should explain the percentage of inertia.

While 21 dimensions were necessary to account for 80% of the explained variability, only 13 dimensions were retained because the other 8 dimensions had eigenvalues below 1%. The 13 dimensions are presented in Figure 2.

**Figure 2:**
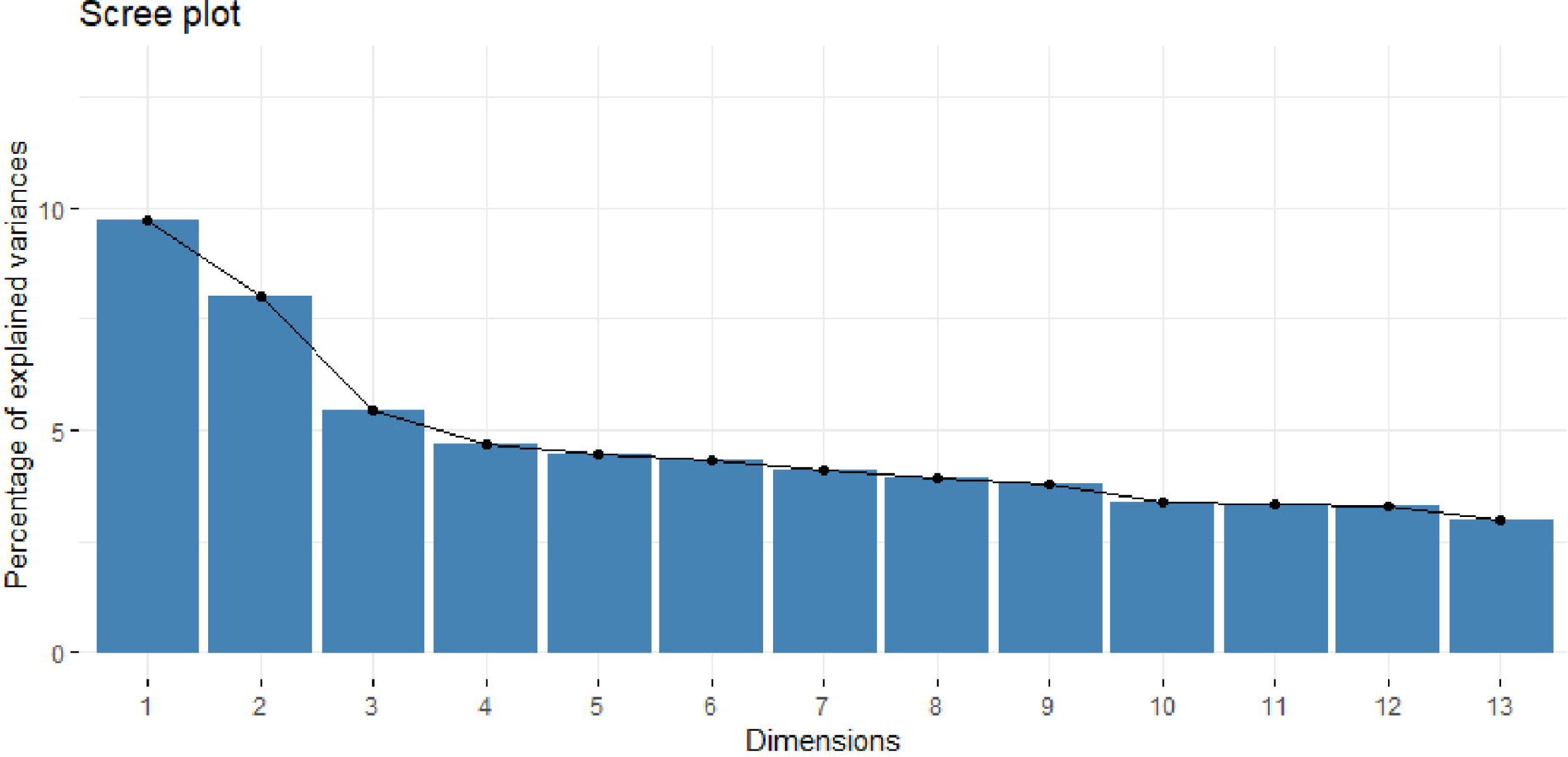
Scree plot representing the percentage of vanance for each of the principal.

### Identification of Three Clusters Using HCPC

The Hierarchical Clustering Post Component Analysis (HCPC) provides a representation of individuals (see figure 3).

**Figure 3:**
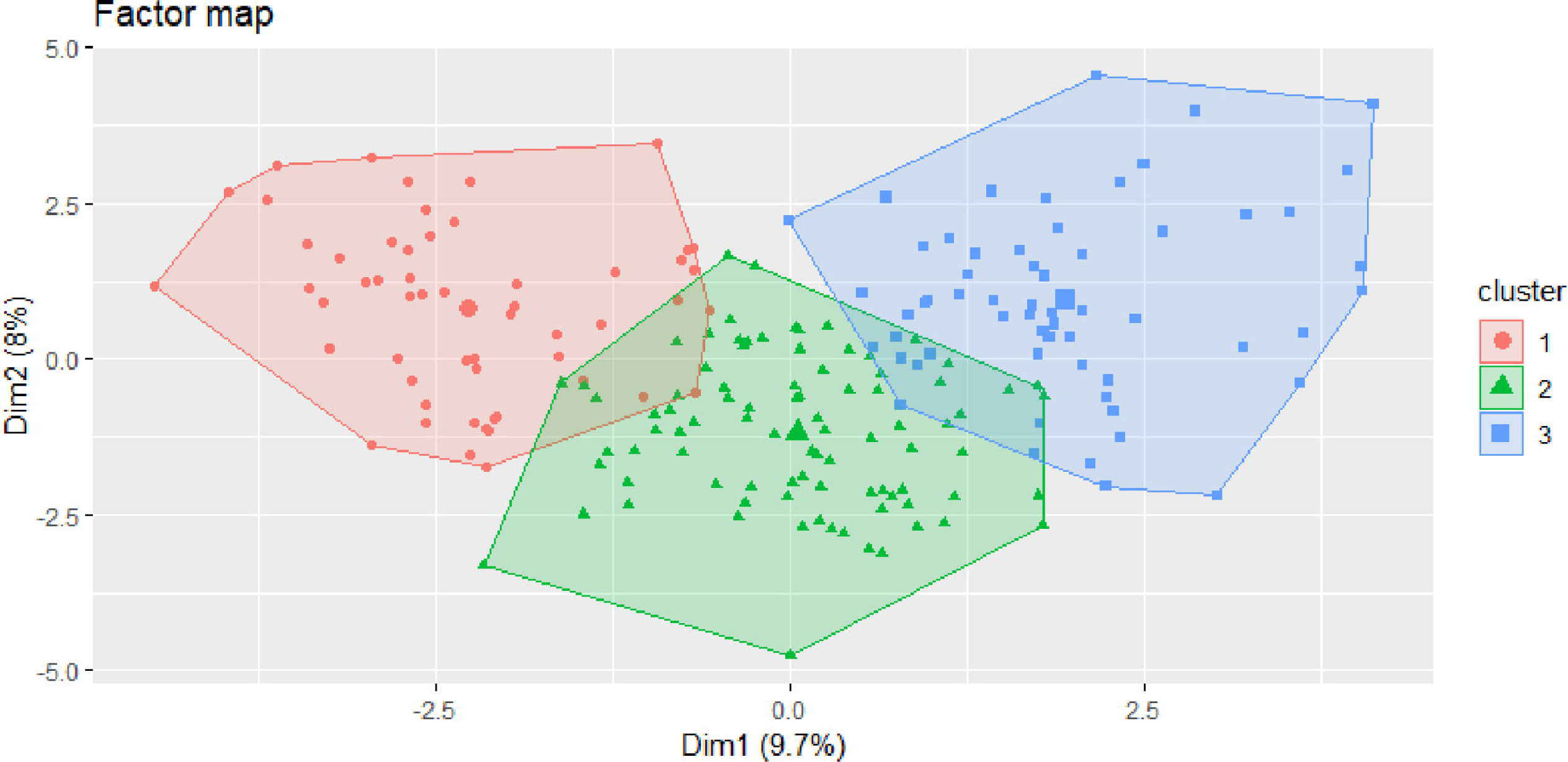
Representation of the clusters on the 2-dimensional factorial plane.

On average, 328 days after SARS-CoV-2 infection, the most distinctive symptoms of long Covid were partitioned into three clusters:

#### Cluster 1 (Pain)

This cluster was characterized by the presence of myalgia. Predominantly male patients and persistent clinical phenotype were represented.

#### Cluster 2 (Central Nervous System)

This cluster primarily featured pure neurocognitive disorders, including concentration, memory, and phasic disorders. It was more prevalent among females and aligned with the long clinical form.

#### Cluster 3 (Anxiety/Depression, Pain, Peripheral Nervous System Disorders with Dysautonomia, Central Nervous System Disorders with Neurocognitive Disorders, Digestive System Disorders)

This cluster exhibited a diverse range of symptoms, including concentration and memory disorders, phasic disorders, headaches, fatigue, anxio-depressive syndrome with sleep and mood disorders, signs of dysautonomia (POTS, dyspnoea, exercise intolerance, palpitations), visual disorders, hearing disorders, taste disorders, paraesthesia, myalgias, joint and chest pain, digestive disorders (nausea, vomiting, abdominal pain), and dizziness.

## Discussion

This study aimed to categorize the various symptoms of long Covid to identify commonalities and differences between individuals, and to subsequently establish a possible connection with clinical forms. Our results show that two of the three clusters identified are linked to the two main clinical forms represented in the sample.

Female sex, a history of psychiatric conditions, and allergies were identified as the primary antecedents of long Covid [3] [17].). Anxiety and depression antecedents were present in 31% and 21% of patients in this cohort, underscoring the interplay between physical and mental aspects in the burden of long Covid, which aligns with the biopsychosocial model [18].

This study’s sample predominantly represented the second wave of Covid-19 infection in Belgium (between April 9, 2020, and February 15, 2021), followed by the first wave (before April 9, 2020). This may suggest that the wild-type strain of SARS-CoV-2 is responsible for long Covid, while the Alpha and Delta variants are primarily associated with the third and fourth waves[19]. However, as patient recruitment is ongoing, it remains possible that patients from subsequent waves will be included, and thus, it cannot be definitively stated that the first two waves are the primary contributors to long Covid.

The most common symptoms of long Covid in this cohort, on average 328 days after the onset of SARS-CoV-2 infection, included neurological disorders (especially concentration and memory issues), fatigue, and sleep disorders. These results are consistent with those reported in Germany, where these symptoms were identified six to twelve months after infection [20].

Furthermore, elevated levels of C-reactive protein were found in long Covid patients compared to those without long Covid who had experienced SARS-CoV-2 infection, based on a meta-analysis of over 20 inflammatory and vascular biomarkers [21]. These findings align with the results of this study, where C-reactive protein was elevated in some patients.

In terms of clustering, few studies have been conducted, each with different patient profiles (hospitalized versus non-hospitalized populations), varying study periods, and different methodologies.

In Dublin, Sweden, they performed multiple correspondence analysis followed by hierarchical ascending classification, identifying three clusters: cluster 1 (pain, myalgias, headaches), cluster 2 (cardiovascular symptoms), and cluster 3 (paucisymptomatic). However, the inclusion period in their study was within 4 weeks of Covid-19 infection, in contrast to the 47 weeks in this study. The difference in inclusion time may explain the disparity in results, as symptoms in long Covid tend to appear chronologically, particularly central nervous system disorders[14].

Four clusters were identified in the United States, using a supervised analysis method that focused on characterizing post-infection pathologies (137 in total) rather than patient-reported symptoms: cluster 1 (cardiac and renal), cluster 2 (respiratory, sleep, and anxiety), cluster 3 (nervous and musculoskeletal systems), and cluster 4 (respiratory and digestive systems) [22]. The differences between their analyses and our study could be attributed to the grouping of cardiovascular symptoms under "pain" and "dysautonomia" in our study, as well as the fact that echocardiographic examinations did not reveal any cardiac abnormalities. Additionally, the disparities may be due to different analysis methods and the earlier recording of symptoms (30 to 180 days post-infection compared to our study’s 328±176 days).

Also in the United States, they identified three clusters in a cohort of hospitalized patients one year after the diagnosis of Covid-19 [23] : cluster 1 (paucisymptomatic, headache), cluster 2 (many symptoms, including anxiety and depression), and cluster 3 (dyspnoea, headache, and cognitive disorders). Notably, the symptoms in their study were reported by patients through questionnaires, and a clinical examination was not conducted to rule out potential Post Intensive Care Syndrome (PICS), which is common in post-ICU patients. Furthermore, their median age was 65 years (compared to 44.5 years in our study), 64% were men, and 34% had been intubated, putting them at higher risk for PICS. Their hierarchical ascending classification was performed directly on the dataset after eliminating missing values. Despite these differences, some similarities exist with our study: cluster 1 is associated with pain, cluster 2 shares similarities with the third cluster in our study, and cluster 3, characterized by neurocognitive disorders, is akin to the second cluster in our study.

Regarding the mechanisms within the different clusters, cluster 1, characterized by myalgia-type pain, could be likened to fibromyalgia syndrome. Some authors have even described long Covid as fibromyalgia[24]. Proposed mechanisms for fibromyalgia include neuroinflammation and dysautonomia, although dysautonomia was not sufficiently present in this cluster to characterize it. Therefore, neuroinflammation is likely the predominant mechanism in this group of patients.

Cluster 2, characterized by pure neurocognitive disorders (concentration, memory, and phasic disorders), might be a consequence of cerebral hypoxia, as this has been suggested as a pathophysiological mechanism for neurological symptoms associated with long Covid[25] [26]. In this group, an associated inflammatory component may also play a role in the onset of neurocognitive disorders. In this context, the interferon (IFN)-complement system has been proposed as the inflammatory mechanism underlying age-related neurocognitive disorders [27].

Cluster 3 (anxiety-depressive syndrome, pain, neurocognitive disorders, dysautonomia, peripheral nervous system disorders, and digestive disorders) may be the result of a combination of factors, including endothelial dysfunction, cerebral hypoxia, inflammation, and psychosocial factors. To gain a better understanding of the pathophysiological mechanisms within these clusters, it would have been necessary to measure various unconventional biomarkers to explore different pathways:

- IL-6, IFN, and complement, which can help assess neuroinflammation [21] [27],
- Von Willebrand factor, ADAMTS-13 protein (a disintegrin and metalloproteinase with thrombospondin type 1 motifs, member 13) as biomarkers of endothelial dysfunction [17] [28],
- HUTT (Head-up tilt table) test to explore orthostatic intolerance, providing a more precise evaluation of dysautonomia [29].

Conventional tests may not be able to differentiate between these clusters.

### Study Limitations

This study has encountered limitations, particularly concerning the standardization of biological tests. The data were collected from patient assessments and sometimes did not include all the necessary information. Ideally, a standardized assessment in both Covid and post-Covid situations would have been more accurate. Additionally, for a more representative sample, it would have been necessary to include all clinical forms of long Covid, including hospitalized patients, who represent a significant proportion of individuals affected by this condition. Exploring psychosocial factors would also have been interesting, as the biopsychosocial model is believed to be involved in the pathophysiology of long Covid.

## Conclusion

This research yielded two main findings: the identification of three clusters of long Covid patients, aligning with two principal clinical forms. Usual biological laboratory tests could not differentiate between these clusters. Therefore, clinicians should conduct meticulous clinical examinations and rule out other diseases to identify different clinical forms of long Covid. Further studies will be necessary to understand the pathophysiological mechanisms or genetic differences present in each cluster/clinical form.

## List of abbreviation

CDC: Centres for Disease Control and Prevention
CFQ: Cognitive Failure Questionnaire
CRU: Clinical Research Unit
CTQ: Childhood Trauma Questionnaire
DN4: neurological pain assessment scale
FSS: Fatigue Severity Scale
HADS: Hospital Anxiety and Depression Scale
HCPC: Hierarchical Clustering Post Component analysis
ICU: Intensive Care Unit
FAMD: Factor analysis of mixed data
LDL: Low Density Lipoprotein
MCAS: Mast Cell Activation Syndrome
MOCA: Montreal Cognitive Assessment
PASC: Post Acute Sequalae of SARS CoV-2
PCL-S: Post Traumatic Checklist Scale
PICS: Post Intensive Care Syndrome
POTS: Postural Orthostatic Tachycardia Syndrome
PTSD: Post-traumatic Stress Disorder
SARS: Severe Acute Respiratory Syndrome
SF-36: Short-Form 36 quality of life scale
SSS-8: Eight item Somatic Symptom Scale
STAI Y-B: State Trait Anxiety Inventory form Y-B
VAS: Visual Analog Scale
WHO: World Health Organization

## Data Availability

All data produced in the present study are available upon reasonable request to the authors.

## Acknowledgement

We gratefully acknowledge:

-Ms. Sahli Chaima for her involvement in the recruitment of participants;

-all the participants to the study for donating their time and the clinical research unit team of the Brugmann university Hospital involved in consenting and recruitment of participants.

## Ethical approval

The study was conducted according to the guidelines of the Declaration of Helsinki and was approved by the ethics committee of Brugmann University Hospital (**ref CE 2022/56**).

## Notes

### Competing Interest Statement

The authors have declared no competing interest.

### Funding Statement

This study did not receive any funding.

### Author Declarations

The ethics committee of Brugmann University Hospital (ref CE 2022/56) gave ethical approval of this work.

### Summary of Updates

The version of the manuscript has been revised to update the methodology which was not clear, particularly about the study design.

